# Inter- and intra-chromosomal modulators of the *APOE* ε2 and ε4 effects on the Alzheimer’s disease risk

**DOI:** 10.1101/2022.06.16.22276523

**Authors:** Alireza Nazarian, Ian Philipp, Irina Culminskaya, Liang He, Alexander M. Kulminski

**Affiliations:** Biodemography of Aging Research Unit, Social Science Research Institute, Duke University, Durham, NC, USA

**Keywords:** Dementia, Aging, LD, Cox Regression, Compound Genotype, Genetic Heterogeneity

## Abstract

The mechanisms of incomplete penetrance of risk modifying impacts of apolipoprotein E (*APOE*) ε2 and ε4 alleles on Alzheimer’s disease (AD) have not been fully understood. We performed genome-wide analysis of differences in linkage disequilibrium (LD) patterns between 6136 AD-affected and 10555 AD-unaffected subjects from five independent studies to explore whether the association of the *APOE* ε2 allele (encoded by rs7412 polymorphism) and ε4 allele (encoded by rs429358 polymorphism) with AD was modulated by autosomal polymorphisms. The LD analysis identified 24 (mostly inter-chromosomal) and 57 (primarily intra-chromosomal) autosomal polymorphisms with significant differences in LD with either rs7412 or rs429358, respectively, between AD-affected and AD-unaffected subjects, indicating their potential modulatory roles. Our Cox regression analysis showed that minor alleles of four inter-chromosomal and ten intra-chromosomal polymorphisms exerted significant modulating effects on the ε2- and ε4-associated AD risks, respectively, and identified ε2-independent (rs2884183 polymorphism, 11q22.3) and ε4-independent (rs483082 polymorphism, 19q13.32) associations with AD. Our functional analysis highlighted ε2- and/or ε4-linked processes affecting the lipid and lipoprotein metabolism, and cell junction organization which may contribute to AD pathogenesis. These findings provide insights into the ε2- and ε4-associated mechanisms of AD pathogenesis, underlying their incomplete penetrance.

## Introduction

The apolipoprotein E (*APOE*) gene is the strongest Alzheimer’s disease (AD)-associated genetic factor [1–3], which can explain 13.4% of phenotypic variance and 25.2% of genetic variance of AD [4]. Minor alleles of the exonic single-nucleotide polymorphisms (SNPs) rs429358 and rs7412 in the *APOE* gene encode the ε4 and ε2 alleles, respectively. The ε2 allele is considered as a protective factor against AD, whereas the ε4 allele is advocated to be a major variant predisposing to AD [3,5].

The *APOE* gene encodes a lipoprotein mainly involved in lipid transfer and metabolism. Nevertheless, its functional impacts are not limited to lipid profile alterations and related vasculopathies [6]. The *APOE* involvement in AD pathogenesis has been widely studied, revealing various molecular and biological processes differentially impacted by different *APOE* alleles. For instance, the ε4 allele has been linked to increased production and decreased clearance of β-amyloid, stress-mediated increased tau hyperphosphorylation, accelerated cortical atrophy (e.g., in the medial temporal lobe), baseline neuronal hyperactivity (e.g., in the hippocampus), reduced cerebral glucose metabolism, damaged synaptic structure and function, increased cytoskeletal and mitochondrial dysfunction, and abnormal hippocampal neurogenesis [7].

Despite strong associations between *APOE* and AD, neither the ε2 nor ε4 allele is considered as a causal factor for AD development [5,8–10]. Addressing the mechanisms of actions of the ε2 and ε4 alleles is essential for understanding AD pathogenesis and AD risk assessment. The complex regional interactions and haplotype structures in the *APOE* locus (19q13.3) have been emphasized by a growing body of studies [11–19]. These studies indicate the potential roles of nearby polymorphisms in modulating the impacts of the *APOE* alleles on AD risks in the form of haplotypes and combinations of genotypes (called compound genotypes). The analyses of haplotypes leverage the idea that AD can be affected by haplotypes driven by genetic linkage between nearby SNPs [20]. The functional linkage may drive, however, compound genotypes consisting of not only local but also distant variants [21].

In this study, we used a comprehensive approach to examine intra- (cis-acting) and inter- (trans-acting) chromosomal modulators of the impacts of the *APOE* rs7412 or rs429358 SNPs on the AD risk in the ε4- or ε2-negative sample. We leveraged samples of the AD-affected (N=6136) and unaffected (N=10555) subjects from five studies: (i) to perform a comparative analysis of LD between rs7412 or rs429358 and other autosomal SNPs in the human genome in the AD-affected and unaffected subjects, (ii) to examine AD risks for carriers of compound genotypes comprised of rs7412 or rs429358 and the identified intra- and inter-chromosomal SNPs in LD with them, and (iii) to identify biological functions and diseases enriched by genes harboring these SNPs.

## Methods

### Study Participants

We used data on subjects of European ancestry from (Table S1): three National Institute on Aging (NIA) Alzheimer’s Disease Centers data (ADCs) from the Alzheimer’s Disease Genetics Consortium (ADGC) initiative [22], whole-genome sequencing (WGS) data from the Alzheimer’s Disease Sequencing Project (ADSP-WGS) [23,24], Cardiovascular Health Study (CHS) [25], Framingham Heart Study (FHS) [26,27], and NIA Late-Onset Alzheimer’s Disease Family Based Study (LOAD FBS) [28]. The ADSP-WGS’s subjects who were also present in other datasets were excluded to make datasets independent. The *APOE* genotypes were either directly reported by original studies (ADGC, ADSP-WGS, FHS) or were determined based on the rs429358 and rs7412 genotypes (CHS and LOAD FBS). The diagnoses of AD cases in the five analyzed datasets were mainly based on the neurologic exams [29,30], and the AD status was reported either directly (ADGC, ADSP-WGS, FHS, LOAD FBS) or in the form of ICD-9 (International Classification of Disease codes, ninth revision) codes (CHS).

### Genotype Data and Quality Control (QC)

We used whole-genome sequencing (ADSP-WGS) and genome-wide data from different array-based platforms (ADGC, CHS, FHS, LOAD FBS). SNPs were first imputed to harmonize them across analyzed datasets [31]. Low-quality data were excluded using *PLINK* [32] as follows: 1) SNPs and subjects with missing rates >5%, 2) SNPs with minor allele frequencies (MAF) <5%, 3) SNPs deviated from Hardy-Weinberg with P<1E-06, and 4) SNPs, subjects, and/or families with Mendel error rates >2% (in ADSP-WGS, FHS, and LOAD FBS which include families). In addition, imputed SNPs with r^2^<0.7 were filtered out (ADGC, CHS, FHS, LOAD FBS). Selecting SNPs presented at least in one study resulted in a set of 1,645,025 SNPs for the analysis.

### Two-stage LD Analysis

#### Design

Our analyses were performed separately in stratified samples obtained by dividing each dataset into four groups based on the *APOE* genotypes and AD status. First, we determined ε4-negative (ε2ε2, ε2ε3, and ε3ε3 genotypes) and ε2-negative (ε4ε4, ε3ε4, and ε3ε3 genotypes) subsamples. Then each subsample was divided into AD-affected and unaffected groups (herein referred to as AD and NAD groups, respectively). We evaluated LD between the *APOE* rs7412 or rs429358 SNP and each SNP in the genome in two stages.

#### Stage 1: LD Analysis in Individual and pooled Datasets

We examined LD (i.e., r statistics) using the haplotype-based method [33–35] in each of the four selected subsamples in each dataset individually and combined. The statistically significant LD estimates were determined using a conservative chi-square test *χ*^*2*^*=r*^*2*^*n* [35], where *n* is the number of subjects rather than gametes to address the uncertainty in inferring haplotypes from unphased genetic data [16,18,36,37]. The variances of the r statistics were calculated using the asymptotic variance method detailed in [37]. The LD analysis was performed using *haplo*.*stats* r package [38].

Stage 1 provided two sets of SNPs in LD with the *APOE* SNPs in each subsample. The first set was generated following the discovery-replication strategy (herein referred to as replication set). In this case SNPs were selected if their LD with the *APOE* SNP attained: 1) genome-wide (P < 5E-08) or suggestive-effect (5E-08 ≤ P < 5E-06) significance in any of the five datasets, which was considered as a discovery set, and 2) Bonferroni-adjusted P<0.0125 (=0.05/4, where 4 is the number of potential replication sets) in at least one of the other four datasets [31]. The second set included SNPs in significant LD with the *APOE* SNPs at genome-wide or suggestive significance in the pooled samples of all five datasets that were not in the replication set.

#### Stage 2: Group-Specific LD

We examined whether SNPs identified in stage 1 had group-specific LD by contrasting *r* between pooled AD and NAD groups, *Δr*=*r*_*AD*_-*r*_*NAD*_, using a permutation test [39,40]. Significant *Δr* indicated SNPs in group-specific LD with rs7412 or rs429358. Bonferroni-adjusted thresholds, accounting for the number of tested SNPs, were used to identify significant findings.

### Analysis of the AD risk

For each group-specific SNP, survival-type analysis was performed to examine the impact of a compound genotype variable (CompG) on the AD risk. The CompG included four compound genotypes comprised of rs7412 or rs429358 genotypes and genotypes of a group-specific SNP (Table 1).

**Table 1.**
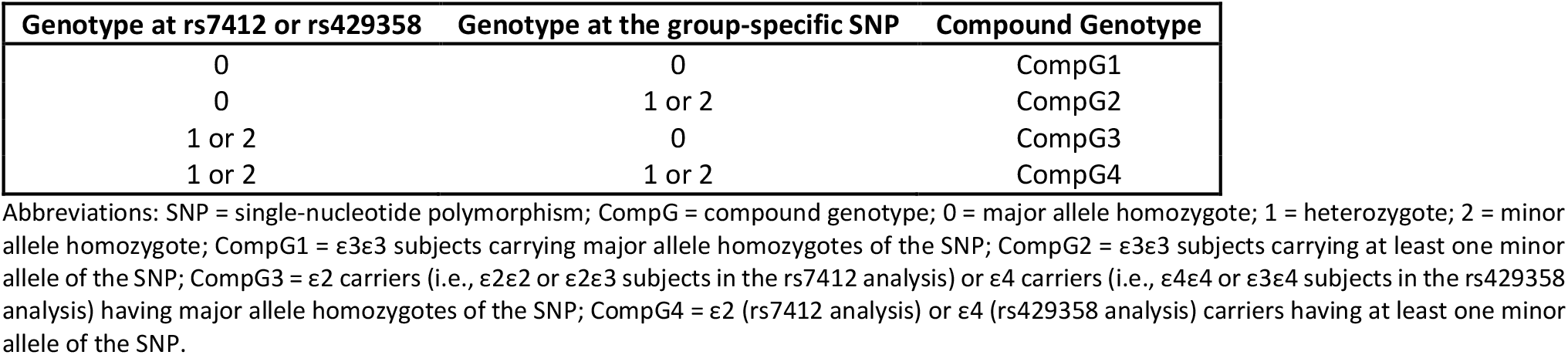
Compound genotype constructed based on the genotypes at rs7412 or rs429358 and the identified group-specific SNPs.

We fitted the Cox regression model (*coxme* and *survival* R packages [41,42]), considering the age at onset of AD as a time variable. We used sex, the top five principal components of genetic data and ADC cohorts (in ADGC) as fixed-effects covariates, and family IDs (LOAD FBS, FHS, ADSP-WGS) as a random-effects covariate. The results from five datasets were combined through inverse-variance meta-analysis using *GWAMA* package [43]. The CompG1 compound genotype was the reference factor level. We used a chi-square test with one degree of freedom [44] to estimate the significance of the difference between the effect sizes for CompG3 and CompG4:

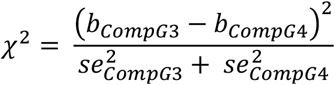

Here, *b*_*CompG3*_ (*se*_*CompG3*_) and *b*_*CompG4*_ (*se*_*compG4*_) are the beta coefficients (standard errors) corresponding to the CompG3 and CompG4 genotypes in the Cox model, respectively. Significant findings were identified at the Bonferroni-adjusted levels correcting for the numbers of ε2- and ε4-associated group-specific SNPs.

### Functional enrichment analysis

*The Database for Annotation, Visualization and Integrated Discovery (DAVID)* [45] and *Metascape* [46] web tools were used to identify gene-enriched *REACTOME* pathways [47] and *DisGeNET* diseases [48]. The analysis was performed for genes harboring SNPs in group-specific LD with rs7412 or rs429358 separately. We used false discovery rate (FDR) adjusted significance cut off at P_FDR_<0.05 [49] to identify significantly enriched terms by two or more genes.

## Results

### SNPs in LD with rs7412 (*APOE* ε2 allele)

In stage 1, we found that 306 SNPs mapped to 27 loci were in LD with rs7412 at P<5E-06 in the AD group (21 SNPs in 9 loci, Table S2), the NAD group (198 SNPs in 20 loci, Table S3), and both AD and NAD groups (87 SNPs, all in the *APOE* locus, Table S4). Of them, we identified LD of rs7412 with 58 SNPs not in the *APOE* locus (or other loci on chromosome 19) in the AD (19 SNPs in 8 loci) or NAD (39 SNPs in 19 loci) groups. For most SNPs, 219 of 306, the magnitudes of LD (i.e., |r|) were smaller in the pooled AD than NAD group (181 of 248 SNPs in the *APOE* locus and 38 of 58 inter-chromosomal SNPs). We also observed that the *r* signs were the same in these two groups for 272 of 306 SNPs.

In stage 2, we found 24 SNPs (Table S5) having group-specific LD with rs7412 at a Bonferroni-adjusted significance P<1.63E-04 (=0.05/306). Of them, 16 SNPs were mapped to 6 non-*APOE* loci. All of them were identified in the pooled sample of either the AD (14 SNPs) or NAD (2 SNPs) group. LD estimates for 14 of these 16 SNPs were characterized by opposite signs of *r* in these groups (Figure 1). Also, 15 of them had larger magnitudes of *r* in the AD group than NAD group. The remaining 8 SNPs were in the *APOE* locus, of which only rs11669338 (*NECTIN2*) attained significance only in NAD group, whereas all the others in both groups. All 8 SNPs had the same signs of *r* in the AD and NAD groups, whose magnitudes were smaller in the AD than NAD group (Figure 1).

**Figure 1.**
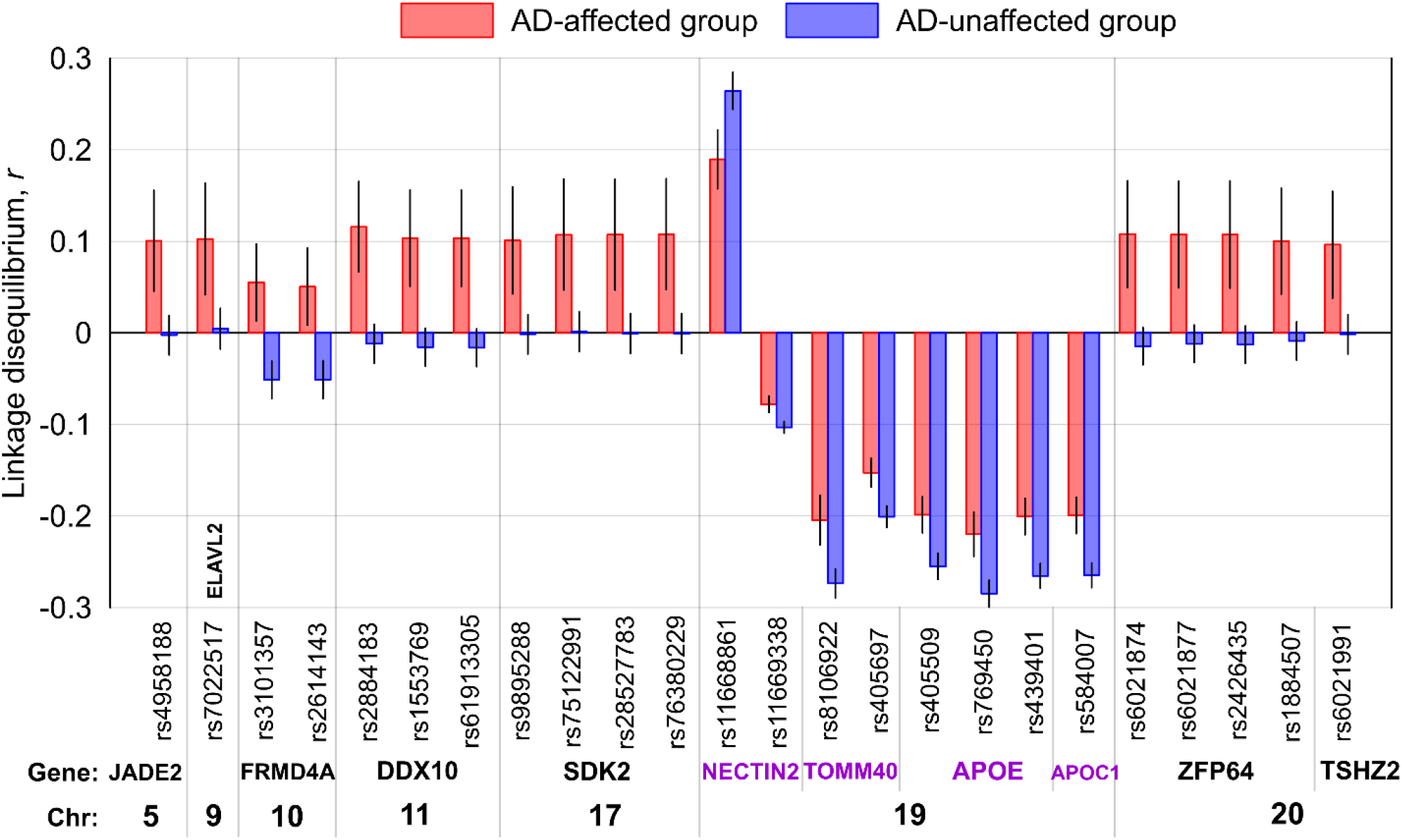
Linkage disequilibrium *r* between the identified group-specific SNPs and rs7412 in the ε4-negative sample of all five datasets combined. The x-axis shows SNP identifiers, genes harboring these SNPs, and chromosomes. Blue boxes: Alzheimer’s disease-affected group (AD). Red boxes: AD-unaffected group (NAD). The vertical lines show 95% confidence intervals.

### SNPs in LD with rs429358 (*APOE* ε4 allele)

In stage 1, we found that rs429358 was in LD with 801 SNPs (143 loci) at P<5E-06 in the AD group (301 SNP in 73 loci, Table S6), the NAD group (351 SNP in 81 loci, Table S7), and both AD and NAD groups (149 SNP; all in the *APOE* locus, except 2 SNPs, Table S8). In the AD and NAD groups, we identified LD of rs429358 with 159 (72 loci) and 344 (80 loci) SNPs not in the *APOE* region, respectively, totaling 503 SNPs. Of all 505 SNPs (154 loci) not in the *APOE* locus in AD, NAD, and AD&NAD groups, one locus harboring *FXYD5* and *FAM187B* genes (11 SNPs, NAD group) was on chromosome 19, and the other 494 SNPs (153 loci) were not on chromosome 19. The LD magnitudes were smaller in the pooled AD than NAD group for 370 of 801 SNPs (270 of 296 SNPs in the *APOE* locus and 161 of 505 SNPs in the non-*APOE* loci). The *r* signs were the same in these two groups for 711 of 801 SNPs.

In stage 2, we identified 57 SNPs with group-specific LD at a Bonferroni-adjusted significance P<6.24E-05 (=0.05/801). As seen in Table S9, 17 of 57 SNPs were mapped to 11 non-*APOE* loci. All of them were identified in the pooled sample of either the AD (10 SNPs) or NAD (7 SNPs) group. The magnitudes of *r* were larger in the pooled AD than NAD sample for SNPs whose significant LD was identified in the AD group and vice versa. The *r* signs for 13 of these 17 SNPs were opposite in these AD and NAD samples. The other 40 SNPs were located in the *APOE* locus. Magnitudes of *r* for all SNPs, except rs769449 (*APOE*), were larger in the pooled AD than NAD sample. For all SNPs, except rs11083767 (*EXOC3L2*), the *r* signs were the same in these AD and NAD samples (Figure 2).

**Figure 2.**
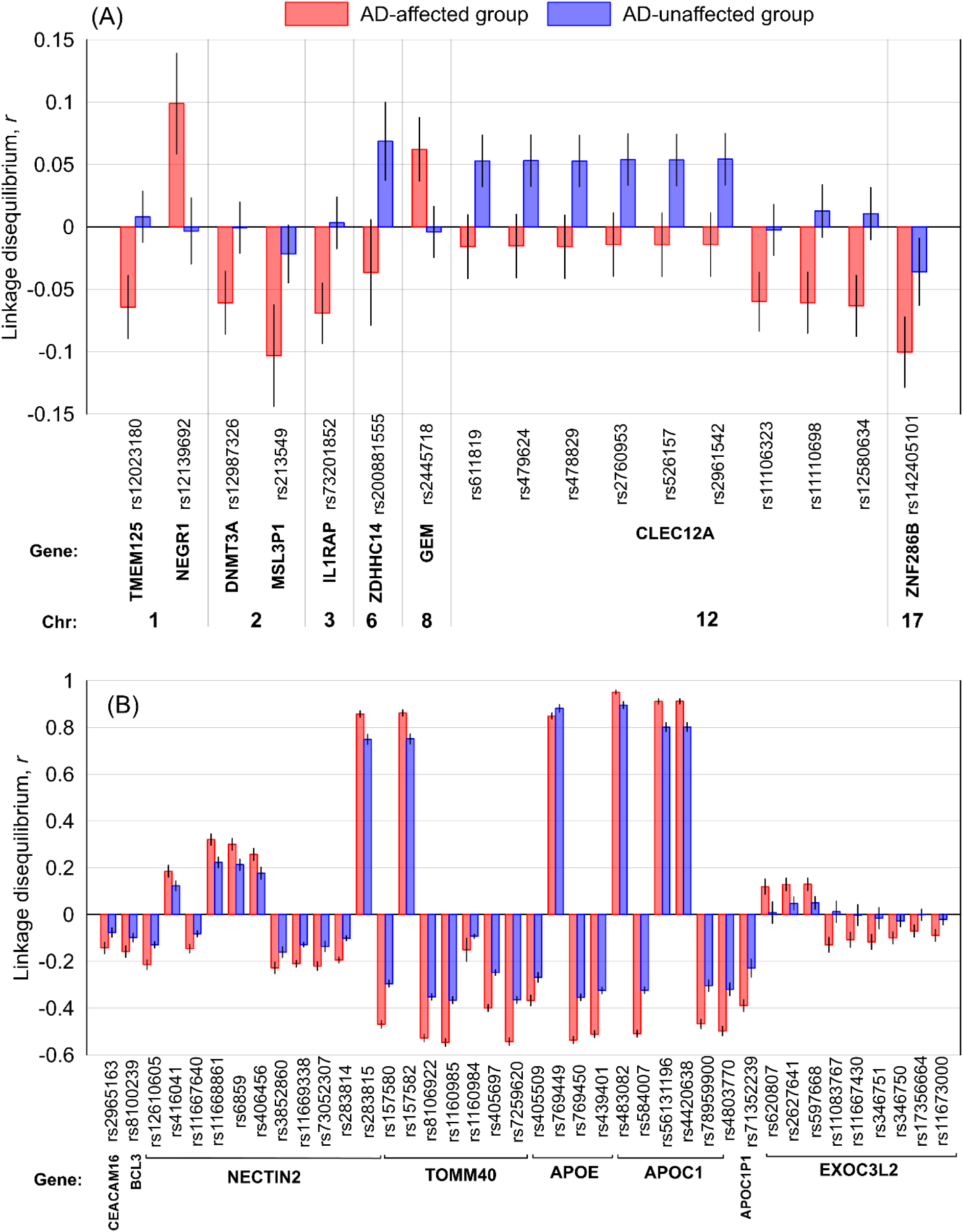
Linkage disequilibrium (LD) *r* between the identified group-specific SNPs and rs429358 in the ε2-negative sample of all five datasets combined. (A) LD for inter-chromosomal SNPs, i.e., SNPs not on chromosome 19. (B) LD for intra-chromosomal SNPs. The x-axis shows SNP identifiers, genes harboring these SNPs, and chromosomes in Figure A. Blue boxes: Alzheimer’s disease-affected group (AD). Red boxes: AD-unaffected group (NAD). The vertical lines show 95% confidence intervals.

### AD risk for carriers of compound genotypes

We performed Cox regression analysis to examine the impact of compound genotypes comprised of a group-specific SNP and either rs7412 (Tables 2 and S10, Figure 3A) or rs429358 (Tables 2 and S11, Figure 3B) on the AD risk. An advantage of using compound genotypes is that we can explicitly examine the effect of a minor allele of a group-specific SNP independently of the effect of the ε2 or ε4 allele (CompG2), the impact of the ε2 or ε4 allele independently of the minor allele of that SNP (CompG3), and combined effects of these minor alleles (CompG4) in the same model with the same reference genotype (CompG1) (Table 1).

**Table 2.**
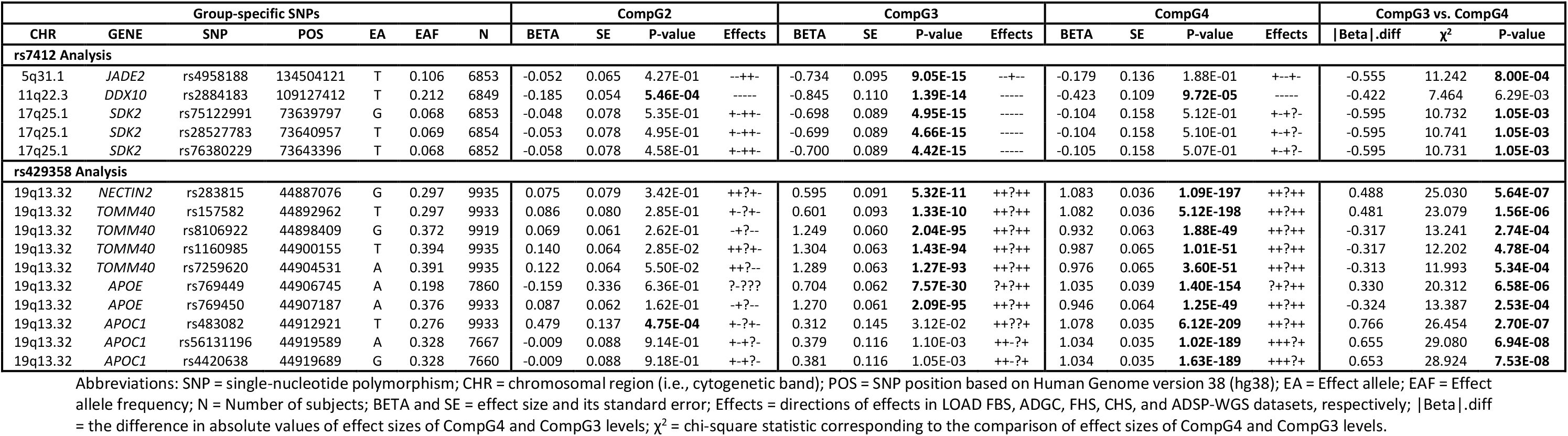
Bonferroni-adjusted significant results from the survival-type meta-analysis of compound genotype (CompG) associations with Alzheimer’s disease risk using SNPs in group-specific LD with rs7412 or rs429358.

**Figure 3.**
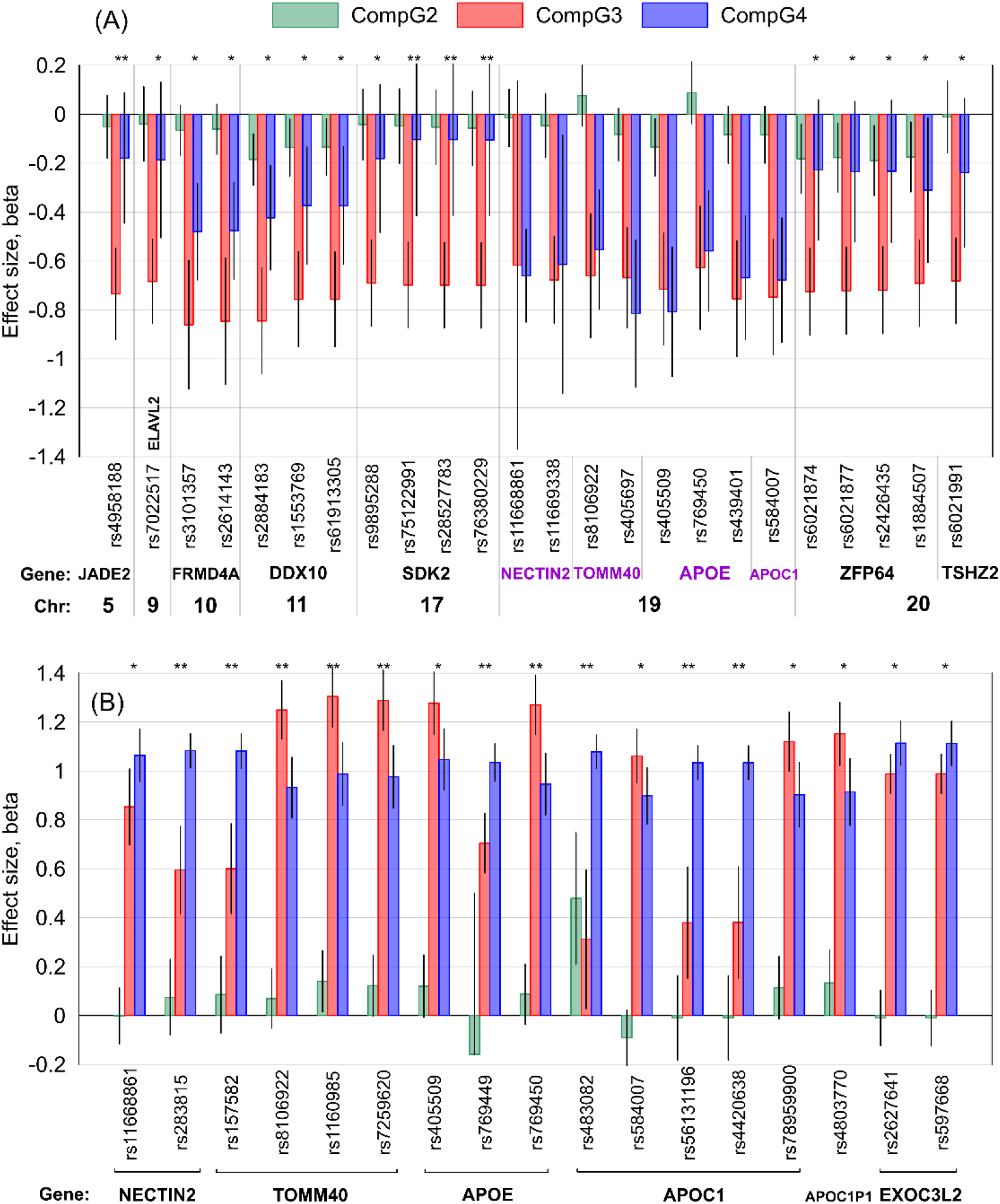
The results of the meta-analysis of the associations of compound genotypes comprised of SNPs (shown on the *x*-axis) in group-specific linkage disequilibrium with (A) rs7412 in the ε4-negative sample or (B) rs429358 in the ε2-negative sample with the Alzheimer’s disease risk. CompG2 (green) indicates ε3ε3 subjects carrying at least one minor allele of the SNP; CompG3 (red) denotes (A) ε2 or (B) ε4 carriers having major allele homozygotes of the SNP; CompG4 (blue) indicates (A) ε2 or (B) ε4 carriers having at least one minor allele of the SNP. CompG1 indicating the ε3ε3 subjects carrying major allele homozygotes of the SNP was the reference. Black vertical lines show 95% confidence intervals (negative direction for rs769449 was truncated for better resolution). The x-axis shows SNP identifiers, genes harboring these SNPs, and chromosomes. One asterisk (*) indicates nominally significant differences in the effects between CompG3 and CompG4 at (A) 2.08E-03≤P<0.05 and (B) 8.77E-04≤P<0.05. Two asterisks (**) indicate Bonferroni-adjusted significance in those differences at (A) P<2.08E-03 and (B) P<8.77E-04. No asterisk indicates non-significant differences in Figure (A). Figure (B) shows only 17 group-specific SNPs for which the differences in the effects between CompG3 and CompG4 attained P<0.05.

#### AD risk for carriers of 24 rs7412-bearing compound genotypes (Tables 2 and S10, Figure 3A)

Our analysis showed that none of eight CompG2 genotypes bearing SNPs from the *APOE* locus attained Bonferroni-adjusted significance P_Bε2_=2.08E-03 (=0.05/24), although rs405509 minor allele was beneficially associated with AD, independently of ε2, at nominal significance P=0.0238. In contrast, six of 16 CompG2 genotypes comprised of rs7412 and non-*APOE* locus SNPs were beneficially associated with AD at the nominal significance (P_Bε2_≤P<0.05). For one CompG2, we observed beneficial association of rs2884183 minor allele (11q22.3, *DDX10*) with AD at P<P_Bε2_ independently of the ε2 allele.

All CompG3 genotypes were beneficially associated with AD (although non-significantly for rs11668861) because of the leading role of the ε2 allele and the lack of minor alleles of the group-specific SNPs. Also, regardless of the significance, all CompG4 genotypes were beneficially associated with AD risk, with 10 of them (seven in the *APOE* locus) reaching P<P_Bε2_. For all 16 group-specific inter-chromosomal SNPs, the effects for CompG4 were smaller in magnitude than those for CompG3 either at the nominal (12 SNPs) or P<P_Bε2_ (four SNPs) significance (Figure 3A).

#### AD risk for carriers of 57 rs429358-bearing compound genotypes (Tables 2 and S11, Figure 3B)

We found that one of 40 intra-chromosomal CompG2 genotypes comprised of rs483082 (*APOC1*) and rs429358 was adversely associated with AD risk independently of the ε4 allele at Bonferroni-adjusted significance P_Bε4_=8.77E-04 (=0.05/57). None of 17 CompG2 with inter-chromosomal SNPs attained P<P_Bε4_.

Each of 57 CompG3 and CompG4 genotypes was adversely associated with the AD risk. None of the differences in the effects between them attained P<P_Bε4_ for inter-chromosomal SNPs. In contrast, we identified seven (P_Bε4_≤P<0.05) and 10 (P<P_Bε4_) differences in the effects between CompG3 and CompG4 for SNPs within the *APOE* locus (Figure 3B).

### Biological functions and diseases

Our analysis was performed for 11 and 19 genes harboring SNPs in group-specific LD with ε2-encoding rs7412 and ε4-encoding rs429358, respectively. We found that 7 and 4 *REACTOME* pathway were enriched at P<0.05 using genes from the ε2 (Figure S1) and ε4 (Figure S2) sets, respectively. Four of them, i.e., “plasma lipoprotein assembly”, “plasma lipoprotein clearance”, “NR1H3 and NR1H2 regulate gene expression linked to cholesterol transport and efflux”, and “NR1H2 and NR1H3-mediated signaling” were enriched in both ε2 and ε4 sets. Three pathways, however, were ε2 specific, including “cell-cell junction organization”, “plasma lipoprotein assembly, remodeling, and clearance”, and “cell junction organization”. There were no enriched ε4 specific pathways.

Disease annotations (Tables S12 and S13) included 14 terms that were enriched at P_FDR_<0.05 by both the ε2 and ε4 gene sets. They were mainly related to neurological diseases (e.g., AD and other dementia phenotypes, memory performance, mild cognitive disorder, and primary progressive aphasia), serum lipid traits (e.g., dyslipoproteinemias, serum Low-density lipoprotein (LDL) cholesterol measurement, and serum total cholesterol measurement), serum albumin measurement, and C-reactive protein measurement.

Seven terms were only enriched in the ε4 set at P_FDR_<0.05 (Table S13) which included mental deterioration, atherogenesis, triglycerides measurement, and high-density lipoprotein measurement as well as multiple hematological and immune system related terms (i.e., autoantibody measurement, acute monocytic leukemia, and peripheral T-cell lymphoma).

## Discussion

Our comprehensive approach examining intra- and inter-chromosomal modulators of the impacts of the *APOE* rs7412 or rs429358 SNP encoding the ε2 or ε4 allele on the AD risk provided four insights.

First, we identified 306 (27 loci) and 801 (143 loci) SNPs in LD with rs7412 and rs429358, respectively, at genome-wide (P<5E-08) or suggestive-effect (5E-08≤P<5E-06) significance in AD, NAD, or both groups. Of them, 58 (27 loci) and 505 (154 loci) SNPs were not on *APOE* locus, indicating potential inter-chromosomal modulators of the impacts of the ε2 or ε4 allele on the AD risk.

Second, among these SNPs, we found significant differences in LD between AD and NAD groups for 24 (16 inter-chromosomal SNPs in 6 loci) and 57 (17 inter-chromosomal SNPs in 11 loci) SNPs with rs7412 and rs429358, respectively, at the Bonferroni-adjusted significance level (Figures 1 and 2, and Tables S5 and S9). This finding strongly supports modulating roles of the intra- and inter-chromosomal SNPs on the impacts of the ε2 or ε4 allele on the AD risk, predominantly tailored to either AD-affected or unaffected subjects.

Third, Cox regression analysis identified Bonferroni-adjusted associations of minor alleles of rs2884183 (11q22.3, *DDX10*) and rs483082 (19q13.32, *APOC1*) with decreased and increased AD risk independently of the ε2 and ε4 alleles, respectively (Table 2).

Fourth, Cox regression analysis revealed that the beneficial and adverse effects of the ε2 and ε4 alleles, respectively, on the AD risks were significantly modulated by other SNPs, and that this modulation was fundamentally different for these alleles. Specifically, the beneficial effect of the ε2 allele was decreased by minor alleles of all 16 group-specific inter-chromosomal SNPs (with a significant decrease at Bonferroni-adjusted level for variants mapped to *JADE2* and *SDK2* genes) (Figure 3A). In contrast, the adverse effect of the ε4 allele was significantly modulated by ten *APOE* locus (intra-chromosomal) SNPs; the ε4 impact was weakened by minor alleles of four SNPs mapped to *TOMM40* and *APOE* genes and major alleles of six SNPs mapped to *NECTIN2, TOMM40, APOE*, and *APOC1* genes (Figure 3B).

The *APOE* locus-specific LD patterns corroborated our previous findings observed for SNP pairs [18] and triples [17]. However, according to the GWAS Catalog [50], none of the identified 33 inter-chromosomal group-specific SNPs have been associated with AD or AD-related pathologies (e.g., amyloid plaque) in previous GWAS at genome-wide or suggestive significance. Rs1884507 (*ZFP64*, in LD with rs7412) and rs12139692 (*NEGR1*, in LD with rs429358) were associated with triglycerides [51] and intelligence [52], respectively, at P<5E-08. Other SNPs mapped to *FRMD4A* [53] and *NEGR1* [54] have been previously associated with AD at genome-wide and suggestive significance, respectively. In addition, several SNPs mapped to the *JADE2, FRMD4A, DDX10, SDK2, ZFP64, TSHZ2, ZDHHC14, NEGR1*, and *SLC5A8* genes, in interaction with SNPs in the other non-*APOE*-locus genes, were associated with AD-related brain pathologies such as diffuse amyloid plaque, PHF-tau, and neurofibrillary tangles at P<5E-08 [55]. Also, an *IL1RAP* variant was previously associated with amyloid plaque accumulation rate at P<5E-08 [56]. Additionally, SNPs mapped to *JADE2, ELAVL2*, and *TSHZ2* have been associated with educational attainment [57] and those mapped to *ELAVL2* and *NEGR1* with intelligence and general cognitive ability [58,59].

Next, we discuss *JADE2* and *SDK2* genes harboring inter-chromosomal SNPs, which significantly modulate the effects of the ε2 allele on AD risk (Table 2). *JADE2* is involved in ubiquitination of histone demethylase *LSD1* [60] and may play roles in the *LSD1*-mediated regulation of neurogenesis and myogenesis [61,62]. *LSD1* is required for neuronal survival and was implicated in tau-induced neurodegeneration in AD and frontotemporal dementia [63,64]. Additionally, *JADE2* (alias *PHF15*) may regulate the microglial inflammatory response [65].

*SDK2* is involved in lamina-specific synaptic connections which are essential to form neuronal circuits in retina that detect motion [66]. Visual impairments including motion detection abnormalities have been reported in AD [67] and Huntington’s disease [68]. Also, visual working memory (i.e., object identification and location recall) were previously associated with the ε4 allele and β-amyloid accumulation [69].

We also highlight *DDX10* gene harboring rs2884183, which is associated with AD risk independently of ε2 (Table 2). The RNA helicase *DDX10* affects ribosome assembly and modulates α-synuclein toxicity [70]. α- Synuclein may synergistically interact with β-amyloid and Tau protein to promote their accumulation [71] and may be involved in the pathogenesis of AD in addition to synucleinopathie (e.g., Parkinson’s disease) [72,73]. *DDX10* may also affect ovarian senescence [74].

Our enrichment analysis of biological functions (Figures S1 and S2) suggested that group-specific LD with rs7412 or rs429358 entails SNPs in genes, which are involved in lipid and lipoprotein metabolism. Additionally, LD with rs7412 entails SNPs in genes, which may contribute to cell junction organization. These biological processes have been implicated in AD pathogenesis [31,75–79]. The disease enrichment analysis (Tables S12 and S13) mostly highlighted the enrichment of AD, dementia phenotypes, and other neurological diseases as well serum lipid traits in both the ε2 and ε4 gene sets. In addition, multiple lipid traits, and neurological and immune system related disorders were enriched in the ε4 gene set.

Investigating the impacts of group-specific SNPs on gene expression revealed that several SNPs in LD with rs7412 (Table S5), including rs11668861 (*NECTIN2*), rs6021874, rs6021877, rs2426435, and rs1884507 (*ZFP64*) are in LD (P<0.0001 in the CEU population of Utah Residents with Northern and Western European Ancestry [80]) with expression quantitative trait loci (eQTLs) whose minor alleles increase *NECTIN2* and *ZFP64* expressions in the brain tissue (Table S14). Also, among SNPs in group-specific LD with rs429358 (Table S9), SNPs mapped to *CLEC12A* (rs611819, rs479624, rs478829, rs2760953, rs526157, and rs2961542) and *NECTIN2* (rs416041, rs11668861, rs6859, rs406456, and rs3852860) are in LD (P<0.0001 [80]) with eQTLs altering the expressions of these two genes. In addition, rs4803770 and rs71352239 (*APOC1P1*) are themselves eQTLs for this gene whose minor alleles decrease *APOC1P1* expression in the brain tissue (Table S14) [81]. In addition, the transcription factor binding sites (TFBS) enrichment [46] shows that that *JADE2, ELAVL2, FRMD4A*, and *APOC1* genes (harboring ε2 group-specific SNPs) have a common TFBS motif corresponding to *RXRB* within ±2kb of their transcription starting sites (P<2.00E-06 and P_FDR_<5.01E-03) [82]. Also, *TMEM125, DNMT3A, ZDHHC14*, and *BCL3* genes (harboring ε4 group-specific SNPs) share a TFBS motif corresponding to *SP3* within ±2kb of their transcription starting sites (P<1.58E-05 and P_FDR_<2.51E-02) [82].

Despite the rigor, this study has limitations. The first is that GWAS datasets do not provide phased genetic data and, therefore, probabilistic estimates of haplotypes may adversely impact the power of LD analyses. Second, due to the small frequency of the ε2 allele in the general population, the LD analysis of rs7412 with the other SNPs may not have optimal statistical power, particularly in the AD-affected group because of the protective role of the ε2 allele against AD. Third, because genotypes were available from WGS in ADSP and genome-wide arrays in the other datasets, we imputed SNPs to harmonize them across all five datasets. Imputation generally results in less accurate genotype calls compared with WGS, particularly in genomic regions with low coverage on the arrays. Low imputation quality may adversely impact the results of the analyses. Although we excluded SNPs with imputation quality of r2 < 0.7 to offset the impacts of potential inaccuracies, replication of the results using directly genotyped SNPs could add robustness to our findings. Fourth, while the Cox regression analysis of genetic associations using AAO of a complex trait provides higher statistical power than the logistic regression analysis of the case-control status [83], we acknowledge limited abilities in determining exact AAO due to slow progression of AD. For instance, AD is not usually diagnosed when the brain pathologies start to develop years before clinical manifestations. Fifth, the small number of genes may affect the accuracy of the functional enrichment analysis. Finally, further stratifying of the AD group based on the pathological information on AD sub-phenotypes would provide valuable insights into the genetic heterogeneity of AD. Also, including subjects with mild cognitive impairment (MCI) in LD analyses as a separate stratum may help to identify APOE allele dependent genetic factors contributing to MCI progression to AD. Such additional stratifications would require large datasets with more comprehensive clinical and pathological data.

## Conclusion

Our comprehensive analysis provides compelling evidence that intra- and inter-chromosomal variants can modulate the impacts of the ε2 and ε4 alleles on the AD risk. The survival-type analysis robustly shows predominant modulating roles of the inter-chromosomal SNPs for the ε2 allele and the *APOE-*region SNPs for the ε4 allele. We identified two variants in *DDX10* (11q22.3) and *APOC1* (19q13.32) genes with beneficial and adverse associations with AD risk independently of the ε2 and ε4 alleles, respectively. Functional enrichment analysis highlighted ε2- and/or ε4-linked processes involved in lipid and lipoprotein metabolism and cell junction organization which have been implicated in AD pathogenesis. Our results advance the understanding of the mechanisms of AD pathogenesis and help improve the accuracy of AD risk assessment.

## Data Availability

Data used in this study can be obtained from dbGaP (https://www.ncbi.nlm.nih.gov/gap/) and NIAGADS (https://www.niagads.org/adsp/content/home).

## Supplementary Information

*Supplementary Information File*: containing *Supporting Acknowledgment*, Table S1, and Figures S1 and S2. Tables S2-S14 in Excel format.

## Acknowledgements

This manuscript was prepared using limited access datasets obtained from dbGaP [accession numbers: phs000372.v1.p1 (ADGC), phs000572.v8.p4 (ADSP), phs000287.v7.p1 (CHS), phs000007.v28.p10 (FHS), and phs000168.v2.p2 (LOAD FBS)] and NIAGADS [accession number: NG00067 (ADSP)]. Please also see the *Supporting Acknowledgment* in the *Supplementary Information* File regarding these five datasets.

## Ethics approval and consent to participate

This study focuses on secondary analysis of data obtained from dbGaP (ADGC, ADSP, CHS, FHS, and LOAD FBS datasets) and NIAGADS (ADSP dataset); and does not involve gathering data from human subjects directly. The data were accessed upon approval by the Duke University Institutional Review Board (IRB) and all analyses were performed following the IRB guidelines.

## References

1. Saunders AM, Strittmatter WJ, Schmechel D, George-Hyslop PH, Pericak-Vance MA, Joo SH, et al. Association of apolipoprotein E allele epsilon 4 with late-onset familial and sporadic Alzheimer’s disease. Neurology. 1993;43:1467–72.

2. Lucotte G, Visvikis S, Leininger-Möler B, David F, Berriche S, Revéilleau S, et al. Association of apolipoprotein E allele ϵ4 with late-onset sporadic Alzheimer’s disease. American Journal of Medical Genetics. 1994;54:286–8.

3. Farrer LA, Cupples LA, Haines JL, Hyman B, Kukull WA, Mayeux R, et al. Effects of age, sex, and ethnicity on the association between apolipoprotein E genotype and Alzheimer disease: a meta-analysis. JAMA. 1997;278:1349–56.

4. Ridge PG, Hoyt KB, Boehme K, Mukherjee S, Crane PK, Haines JL, et al. Assessment of the genetic variance of late-onset Alzheimer’s disease. Neurobiol Aging. 2016;41:200.e13-200.e20.

5. Genin E, Hannequin D, Wallon D, Sleegers K, Hiltunen M, Combarros O, et al. APOE and Alzheimer disease: a major gene with semi-dominant inheritance. Mol Psychiatry. 2011;16:903–7.

6. Belloy ME, Napolioni V, Greicius MD. A quarter century of APOE and Alzheimer’s disease: progress to date and the path Forward. Neuron. 2019;101:820–38.

7. Liu C-C, Liu C-C, Kanekiyo T, Xu H, Bu G. Apolipoprotein E and Alzheimer disease: risk, mechanisms and therapy. Nat Rev Neurol. 2013;9:106–18.

8. Corder EH, Saunders AM, Strittmatter WJ, Schmechel DE, Gaskell PC, Small GW, et al. Gene dose of apolipoprotein E type 4 allele and the risk of Alzheimer’s disease in late onset families. Science. 1993;261:921–3.

9. Freudenberg-Hua Y, Freudenberg J, Vacic V, Abhyankar A, Emde A-K, Ben-Avraham D, et al. Disease variants in genomes of 44 centenarians. Mol Genet Genomic Med. 2014;2:438–50.

10. Kulminski AM, Philipp I, Loika Y, He L, Culminskaya I. Protective association of the ε2/ε3 heterozygote with Alzheimer’s disease is strengthened by TOMM40-APOE variants in men. Alzheimers Dement. 2021;

11. Templeton AR, Maxwell T, Posada D, Stengård JH, Boerwinkle E, Sing CF. Tree scanning: a method for using haplotype trees in phenotype/genotype association studies. Genetics. 2005;169:441–53.

12. Yu C-E, Seltman H, Peskind ER, Galloway N, Zhou PX, Rosenthal E, et al. Comprehensive analysis of APOE and selected proximate markers for late-onset Alzheimer’s disease: patterns of linkage disequilibrium and disease/marker association. Genomics. 2007;89:655–65.

13. Lescai F, Chiamenti AM, Codemo A, Pirazzini C, D’Agostino G, Ruaro C, et al. An APOE haplotype associated with decreased ε4 expression increases the risk of late onset Alzheimer’s disease. Journal of Alzheimer’s Disease. IOS Press; 2011;24:235–45.

14. Lutz MW, Crenshaw D, Welsh-Bohmer KA, Burns DK, Roses AD. New genetic approaches to AD: lessons from APOE-TOMM40 phylogenetics. Curr Neurol Neurosci Rep. 2016;16:48.

15. Babenko VN, Afonnikov DA, Ignatieva EV, Klimov AV, Gusev FE, Rogaev EI. Haplotype analysis of APOE intragenic SNPs. BMC Neurosci. 2018;19:16.

16. Kulminski AM, Huang J, Wang J, He L, Loika Y, Culminskaya I. Apolipoprotein E region molecular signatures of Alzheimer’s disease. Aging Cell. 2018;e12779.

17. Kulminski AM, Philipp I, Loika Y, He L, Culminskaya I. Haplotype architecture of the Alzheimer’s risk in the APOE region via co-skewness. Alzheimers Dement (Amst). 2020;12:e12129.

18. Kulminski AM, Shu L, Loika Y, Nazarian A, Arbeev K, Ukraintseva S, et al. APOE region molecular signatures of Alzheimer’s disease across races/ethnicities. Neurobiology of Aging. 2020;87:141.e1-141.e8.

19. Zhou X, Chen Y, Mok KY, Kwok TCY, Mok VCT, Guo Q, et al. Non-coding variability at the APOE locus contributes to the Alzheimer’s risk. Nat Commun. 2019;10:3310.

20. Roses AD, Lutz MW, Amrine-Madsen H, Saunders AM, Crenshaw DG, Sundseth SS, et al. A TOMM40 variable-length polymorphism predicts the age of late-onset Alzheimer’s disease. Pharmacogenomics J. 2010;10:375–84.

21. Linghu B, Franzosa EA, Xia Y. Construction of Functional Linkage Gene Networks by Data Integration. In: Mamitsuka H, DeLisi C, Kanehisa M, editors. Data Mining for Systems Biology: Methods and Protocols [Internet]. Totowa, NJ: Humana Press; 2013 [cited 2021 Jul 8]. p. 215–32. Available from: https://doi.org/10.1007/978-1-62703-107-3_14

22. Naj AC, Jun G, Beecham GW, Wang L-S, Vardarajan BN, Buros J, et al. Common variants at MS4A4/MS4A6E, CD2AP, CD33 and EPHA1 are associated with late-onset Alzheimer’s disease. Nat Genet. 2011;43:436–41.

23. Beecham GW, Bis JC, Martin ER, Choi S-H, DeStefano AL, van Duijn CM, et al. The Alzheimer’s Disease Sequencing Project: study design and sample selection. Neurol Genet. 2017;3:e194.

24. Crane PK, Foroud T, Montine TJ, Larson EB. Alzheimer’s Disease Sequencing Project discovery and replication criteria for cases and controls: Data from a community-based prospective cohort study with autopsy follow-up. Alzheimers Dement. 2017;13:1410–3.

25. Fried LP, Borhani NO, Enright P, Furberg CD, Gardin JM, Kronmal RA, et al. The cardiovascular health study: design and rationale. Ann Epidemiol. 1991;1:263–76.

26. Dawber TR, Meadors GF, Moore FE. Epidemiological approaches to heart disease: the Framingham study. Am J Public Health Nations Health. 1951;41:279–86.

27. Feinleib M, Kannel WB, Garrison RJ, McNamara PM, Castelli WP. The Framingham offspring study: design and preliminary data. Prev Med. 1975;4:518–25.

28. Lee JH, Cheng R, Graff-Radford N, Foroud T, Mayeux R. Analyses of the national institute on aging late-onset Alzheimer’s disease family study: implication of additional loci. Arch Neurol. 2008;65:1518–26.

29. McKhann G, Drachman D, Folstein M, Katzman R, Price D, Stadlan EM. Clinical diagnosis of Alzheimer’s disease: report of the NINCDS-ADRDA Work Group under the auspices of Department of Health and Human Services Task Force on Alzheimer’s Disease. Neurology. 1984;34:939–44.

30. McKhann GM, Knopman DS, Chertkow H, Hyman BT, Jack CR, Kawas CH, et al. The diagnosis of dementia due to Alzheimer’s disease: recommendations from the National Institute on Aging-Alzheimer’s Association workgroups on diagnostic guidelines for Alzheimer’s disease. Alzheimer’s & Dementia. 2011;7:263–9.

31. Nazarian A, Yashin AI, Kulminski AM. Genome-wide analysis of genetic predisposition to Alzheimer’s disease and related sex disparities. Alzheimer’s Research & Therapy. 2019;11:5.

32. Purcell S, Neale B, Todd-Brown K, Thomas L, Ferreira MAR, Bender D, et al. PLINK: a tool set for whole-genome association and population-based linkage analyses. Am J Hum Genet. 2007;81:559–75.

33. Weir BS. Inferences about linkage disequilibrium. Biometrics. [Wiley, International Biometric Society]; 1979;35:235–54.

34. Weir BS, Cockerham CC. Estimation of linkage disequilibrium in randomly mating populations. Heredity. Nature Publishing Group; 1979;42:105–11.

35. Lewontin RC. On measures of gametic disequilibrium. Genetics. 1988;120:849–52.

36. Zaykin DV, Meng Z, Ehm MG. Contrasting linkage-disequilibrium patterns between cases and controls as a novel association-mapping method. Am J Hum Genet. 2006;78:737–46.

37. Wellek S, Ziegler A. A genotype-based approach to assessing the association between single nucleotide polymorphisms. Hum Hered. 2009;67:128–39.

38. Sinnwell J, Schaid D. haplo.stats: statistical analysis of haplotypes with traits and covariates when linkage phase is ambiguous [Internet]. 2020 [cited 2021 Jun 28]. Available from: https://CRAN.R-project.org/package=haplo.stats

39. Krzanowski WJ. Permutational tests for correlation matrices. Stat Comput. 1993;3:37–44.

40. Kulminski AM, Huang J, Wang J, He L, Loika Y, Culminskaya I. Apolipoprotein E region molecular signatures of Alzheimer’s disease. Aging Cell. 2018;17:e12779.

41. Therneau TM. coxme: a package for mixed effects cox models in R. R package version 2.2-16 [Internet]. 2020 [cited 2021 Aug 15]. Available from: https://CRAN.R-project.org/package=coxme

42. Therneau TM. survival: a package for survival analysis in R. R package version 3.2-13 [Internet]. 2021 [cited 2021 Aug 15]. Available from: https://CRAN.R-project.org/package=survival

43. Mägi R, Morris AP. GWAMA: software for genome-wide association meta-analysis. BMC Bioinformatics. 2010;11:288.

44. Allison PD. Comparing logit and probit coefficients across groups. Sociological Methods & Research. 1999;28:186–208.

45. Sherman BT, Hao M, Qiu J, Jiao X, Baseler MW, Lane HC, et al. DAVID: a web server for functional enrichment analysis and functional annotation of gene lists (2021 update). Nucleic Acids Research. 2022;gkac194.

46. Zhou Y, Zhou B, Pache L, Chang M, Khodabakhshi AH, Tanaseichuk O, et al. Metascape provides a biologist-oriented resource for the analysis of systems-level datasets. Nat Commun. 2019;10:1523.

47. Fabregat A, Jupe S, Matthews L, Sidiropoulos K, Gillespie M, Garapati P, et al. The reactome pathway knowledgebase. Nucleic Acids Res. 2018;46:D649–55.

48. Piñero J, Bravo À, Queralt-Rosinach N, Gutiérrez-Sacristán A, Deu-Pons J, Centeno E, et al. DisGeNET: a comprehensive platform integrating information on human disease-associated genes and variants. Nucleic Acids Res. 2017;45:D833–9.

49. Benjamini Y, Hochberg Y. Controlling the false discovery rate: a practical and powerful approach to multiple testing. Journal of the Royal Statistical Society Series B (Methodological). 1995;57:289–300.

50. MacArthur J, Bowler E, Cerezo M, Gil L, Hall P, Hastings E, et al. The new NHGRI-EBI Catalog of published genome-wide association studies (GWAS Catalog). Nucleic Acids Res. 2017;45:D896–901.

51. Klarin D, Damrauer SM, Cho K, Sun YV, Teslovich TM, Honerlaw J, et al. Genetics of blood lipids among ∼300,000 multi-ethnic participants of the Million Veteran Program. Nat Genet. 2018;50:1514–23.

52. Hill WD, Marioni RE, Maghzian O, Ritchie SJ, Hagenaars SP, McIntosh AM, et al. A combined analysis of genetically correlated traits identifies 187 loci and a role for neurogenesis and myelination in intelligence. Mol Psychiatry. 2019;24:169–81.

53. Lambert J-C, Grenier-Boley B, Harold D, Zelenika D, Chouraki V, Kamatani Y, et al. Genome-wide haplotype association study identifies the FRMD4A gene as a risk locus for Alzheimer’s disease. Mol Psychiatry. 2013;18:461–70.

54. Sherva R, Gross A, Mukherjee S, Koesterer R, Amouyel P, Bellenguez C, et al. Genome-wide association study of rate of cognitive decline in Alzheimer’s disease patients identifies novel genes and pathways. Alzheimers Dement. 2020;16:1134–45.

55. Wang H, Yang J, Schneider JA, De Jager PL, Bennett DA, Zhang H-Y. Genome-wide interaction analysis of pathological hallmarks in Alzheimer’s disease. Neurobiol Aging. 2020;93:61–8.

56. Ramanan VK, Risacher SL, Nho K, Kim S, Shen L, McDonald BC, et al. GWAS of longitudinal amyloid accumulation on 18F-florbetapir PET in Alzheimer’s disease implicates microglial activation gene IL1RAP. Brain. 2015;138:3076–88.

57. Lee JJ, Wedow R, Okbay A, Kong E, Maghzian O, Zacher M, et al. Gene discovery and polygenic prediction from a genome-wide association study of educational attainment in 1.1 million individuals. Nat Genet. 2018;50:1112–21.

58. Davies G, Lam M, Harris SE, Trampush JW, Luciano M, Hill WD, et al. Study of 300,486 individuals identifies 148 independent genetic loci influencing general cognitive function. Nat Commun. 2018;9:2098.

59. Savage JE, Jansen PR, Stringer S, Watanabe K, Bryois J, de Leeuw CA, et al. Genome-wide association meta-analysis in 269,867 individuals identifies new genetic and functional links to intelligence. Nat Genet. 2018;50:912–9.

60. Maiques-Diaz A, Somervaille TC. LSD1: biologic roles and therapeutic targeting. Epigenomics. 2016;8:1103–16.

61. Han X, Gui B, Xiong C, Zhao L, Liang J, Sun L, et al. Destabilizing LSD1 by Jade-2 promotes neurogenesis: an antibraking system in neural development. Mol Cell. 2014;55:482–94.

62. Anan K, Hino S, Shimizu N, Sakamoto A, Nagaoka K, Takase R, et al. LSD1 mediates metabolic reprogramming by glucocorticoids during myogenic differentiation. Nucleic Acids Res. 2018;46:5441–54.

63. Christopher MA, Myrick DA, Barwick BG, Engstrom AK, Porter-Stransky KA, Boss JM, et al. LSD1 protects against hippocampal and cortical neurodegeneration. Nat Commun. 2017;8:805.

64. Engstrom AK, Walker AC, Moudgal RA, Myrick DA, Kyle SM, Bai Y, et al. The inhibition of LSD1 via sequestration contributes to tau-mediated neurodegeneration. Proc Natl Acad Sci U S A. 2020;117:29133–43.

65. Muroy SE, Timblin GA, Preininger MK, Cedillo P, Saijo K. Phf15 - a novel transcriptional repressor regulating inflammation in a mouse microglial cell line. Neuroimmunology and Neuroinflammation. OAE Publishing Inc.; 2020;7:166–82.

66. Krishnaswamy A, Yamagata M, Duan X, Hong YK, Sanes JR. Sidekick 2 directs formation of a retinal circuit that detects differential motion. Nature. 2015;524:466–70.

67. Cunha JP, Moura-Coelho N, Proença RP, Dias-Santos A, Ferreira J, Louro C, et al. Alzheimer’s disease: A review of its visual system neuropathology. Optical coherence tomography-a potential role as a study tool in vivo. Graefes Arch Clin Exp Ophthalmol. 2016;254:2079–92.

68. Muratori LM, Evinger L, Reilmann R. F3 Biological motion perception in huntington’s disease. J Neurol Neurosurg Psychiatry. BMJ Publishing Group Ltd; 2016;87:A49–A49.

69. Lu K, Nicholas JM, Pertzov Y, Grogan J, Husain M, Pavisic IM, et al. Dissociable effects of APOE ε4 and β-amyloid pathology on visual working memory. Nat Aging. 2021;1:1002–9.

70. Popova B, Wang D, Pätz C, Akkermann D, Lázaro DF, Galka D, et al. DEAD-box RNA helicase Dbp4/DDX10 is an enhancer of α-synuclein toxicity and oligomerization. PLoS Genet. 2021;17:e1009407.

71. Clinton LK, Blurton-Jones M, Myczek K, Trojanowski JQ, LaFerla FM. Synergistic Interactions between Abeta, tau, and alpha-synuclein: acceleration of neuropathology and cognitive decline. J Neurosci. 2010;30:7281–9.

72. Twohig D, Nielsen HM. α-synuclein in the pathophysiology of Alzheimer’s disease. Mol Neurodegener. 2019;14:23.

73. Monge-García V, García-Ayllón M-S, Sáez-Valero J, Sánchez-Payá J, Navarrete-Rueda F, Manzanares-Robles J, et al. Relation between alpha-synuclein and core CSF biomarkers of Alzheimer’s disease. Medicina (Kaunas). 2021;57:954.

74. Cai H, Chang T, Li Y, Jia Y, Li H, Zhang M, et al. Circular DDX10 is associated with ovarian function and assisted reproductive technology outcomes through modulating the proliferation and steroidogenesis of granulosa cells. Aging (Albany NY). 2021;13:9592–612.

75. Stamatovic SM, Keep RF, Andjelkovic AV. Brain endothelial cell-cell junctions: how to “open” the blood brain barrier. Curr Neuropharmacol. 2008;6:179–92.

76. Stamatovic SM, Johnson AM, Keep RF, Andjelkovic AV. Junctional proteins of the blood-brain barrier: New insights into function and dysfunction. Tissue Barriers [Internet]. 2016 [cited 2019 Apr 11];4. Available from: https://www.ncbi.nlm.nih.gov/pmc/articles/PMC4836471/

77. Liu Q, Zhang J. Lipid metabolism in Alzheimer’s disease. Neurosci Bull. 2014;30:331–45.

78. Nazarian A, Arbeev KG, Yashkin AP, Kulminski AM. Genetic heterogeneity of Alzheimer’s disease in subjects with and without hypertension. GeroScience. 2019;41:137–54.

79. Leshchyns’ka I, Sytnyk V. Synaptic cell adhesion molecules in Alzheimer’s disease. Neural Plast. 2016;2016.

80. Machiela MJ, Chanock SJ. LDlink: a web-based application for exploring population-specific haplotype structure and linking correlated alleles of possible functional variants. Bioinformatics. 2015;31:3555–7.

81. GTEx Consortium. Genetic effects on gene expression across human tissues. Nature. 2017;550:204–13.

82. Subramanian A, Tamayo P, Mootha VK, Mukherjee S, Ebert BL, Gillette MA, et al. Gene set enrichment analysis: A knowledge-based approach for interpreting genome-wide expression profiles. PNAS. 2005;102:15545–50.

83. He L, Kulminski AM. Fast algorithms for conducting large-scale GWAS of age-at-onset traits using cox mixed-effects models. Genetics. 2020;215:41–58.

