## Supplementary Information File for "Inter- and intra-chromosomal modulators of the *APOE* ε2 and ε4 effects on the Alzheimer’s disease risk"

**This file includes:**

Supporting Acknowledgment text

Table S1

Figures S1 and S2

**Other Supplementary Materials for this manuscript include Tables S2-S14 in Excel format and provided as separate files:**

Table S2: SNPs in LD with rs7412 identified only in the AD-affected group.

Table S3: SNPs in LD with rs7412 identified only in the AD-unaffected group.

Table S4: SNPs in LD with rs7412 identified in both AD-affected and unaffected groups.

Table S5: SNPs in group-specific LD with rs7412.

Table S6: SNPs in LD with rs429358 identified only in the AD-affected group.

Table S7: SNPs in LD with rs429358 identified only in the AD-unaffected group.

Table S8: SNPs in LD with rs429358 identified in both AD-affected and unaffected groups.

Table S9: SNPs in group-specific LD with rs429358.

Table S10: survival-type meta-analysis of compound genotype associations with AD risk using SNPs in group-specific LD with rs7412.

Table S11: survival-type meta-analysis of compound genotype associations with AD risk using SNPs in group-specific LD with rs429358.

Table S12: Disease enrichment analysis of the genes harboring SNPs in group-specific LD with rs7412.

Table S13: Disease enrichment analysis of the genes harboring SNPs in group-specific LD with rs429358.

Table S14: eQTLs in LD with group-specific SNPs.

**Supporting Acknowledgment**

This research was supported by Grants from the National Institute on Aging (R01AG047310, R01AG061853, R01AG065477, R01AG070488, and P01AG043352). The funders had no role in study design, data collection and analysis, decision to publish, or manuscript preparation. The content is solely the responsibility of the authors and does not necessarily represent the official views of the National Institutes of Health.

This manuscript was prepared using limited access datasets obtained from dbGaP [accession numbers: phs000372.v1.p1 (ADGC), phs000572.v8.p4 (ADSP), phs000287.v7.p1 (CHS), phs000007.v28.p10 (FHS), and phs000168.v2.p2 (LOAD FBS)] and NIAGADS [accession number: NG00067 (ADSP)].

**LOAD FBS**

Funding support for the Late Onset Alzheimer's Disease Family Study (LOAD FBS) was provided through the Division of Neuroscience, NIA. The LOAD FBS includes a genome-wide association study funded as part of the Division of Neuroscience, NIA. Assistance with phenotype harmonization and genotype cleaning, as well as with general study coordination, was provided by Genetic Consortium for Late Onset Alzheimer's Disease. This manuscript was not prepared in collaboration with LOAD FBS investigators and does not necessarily reflect the opinions or views of LOAD FBS.

**FHS**

The Framingham Heart Study (FHS) is conducted and supported by the National Heart, Lung, and Blood Institute (NHLBI) in collaboration with Boston University (Contract No. N01-HC-25195 and HHSN268201500001I). This manuscript was not prepared in collaboration with investigators of the FHS and does not necessarily reflect the opinions or views of the FHS, Boston University, or NHLBI. Funding for SHARe Affymetrix genotyping was provided by NHLBI Contract N02-HL-64278. SHARe Illumina genotyping was provided under an agreement between Illumina and Boston University. Funding for CARe genotyping was provided by NHLBI Contract N01-HC-65226. Funding support for the Framingham Dementia dataset was provided by NIH/NIA grant R01 AG08122. Funding support for the Framingham Inflammatory Markers was provided by NIH grants R01 HL064753, R01 HL076784 and R01 AG028321. Funding support for the Framingham C-reactive Protein dataset was provided by NIH grants R01 HL064753, R01 HL076784 and R01 AG028321. Funding support for the Framingham Adiponectin dataset was provided by NIH/NHLBI grant R01-DK-080739. Funding support for the Framingham Interleukin-6 dataset was provided by NIH grants R01 HL064753, R01 HL076784 and R01 AG028321.

**CHS**

The Cardiovascular Health Study (CHS) was supported by contracts HHSN268201200036C, HHSN268200800007C, N01-HC-85079, N01-HC-85080, N01-HC-85081, N01-HC-85082, N01-HC-85083, N01-HC-85084, N01-HC-85085, N01-HC-85086, N01-HC-35129, N01 HC-15103, N01 HC-55222, N01-HC-75150, N01-HC-45133, and N01-HC-85239; grant numbers U01 HL080295 and U01 HL130014 from the National Heart, Lung, and Blood Institute (NHLBI), and R01 AG-023629 from the National Institute on Aging, with additional contribution from the National Institute of Neurological Disorders and Stroke. A full list of principal CHS investigators and institutions can be found at https://chs-nhlbi.org/pi. This manuscript was not prepared in collaboration with CHS investigators and does not necessarily reflect the opinions or views of CHS, or the NHLBI. Support for the genotyping through the CARe Study was provided by NHLBI Contract N01-HC-65226. Additional support for infrastructure was provided by HL105756 and additional genotyping among the African-American cohort was supported in part by HL085251. DNA handling and genotyping at Cedars-Sinai Medical Center was supported in part by National Center for Research Resources grant UL1RR033176, now at the National Center for Advancing Translational Technologies CTSI grant UL1TR000124; in addition to the National Institute of Diabetes and Digestive and Kidney Diseases grant DK063491 to the Southern California Diabetes Endocrinology Research Center.

**ADGC**

Funding support for the Alzheimer’s Disease Genetics Consortium (ADGC) was provided through the NIA Division of Neuroscience (U01-AG032984).

**ADSP**

The Alzheimer’s Disease Sequencing Project (ADSP) is comprised of two Alzheimer’s Disease (AD) genetics consortia and three National Human Genome Research Institute (NHGRI) funded Large Scale Sequencing and Analysis Centers (LSAC). The two AD genetics consortia are the Alzheimer’s Disease Genetics Consortium (ADGC) funded by NIA (U01 AG032984), and the Cohorts for Heart and Aging Research in Genomic Epidemiology (CHARGE) funded by NIA (R01 AG033193), the National Heart, Lung, and Blood Institute (NHLBI), other National Institute of Health (NIH) institutes and other foreign governmental and non-governmental organizations. The Discovery Phase analysis of sequence data is supported through UF1AG047133 (to Drs. Schellenberg, Farrer, Pericak-Vance, Mayeux, and Haines); U01AG049505 to Dr. Seshadri; U01AG049506 to Dr. Boerwinkle; U01AG049507 to Dr. Wijsman; and U01AG049508 to Dr. Goate and the Discovery Extension Phase analysis is supported through U01AG052411 to Dr. Goate, U01AG052410 to Dr. Pericak-Vance and U01 AG052409 to Drs. Seshadri and Fornage.

Sequencing for the Follow Up Study (FUS) is supported through U01AG057659 (to Drs. PericakVance, Mayeux, and Vardarajan) and U01AG062943 (to Drs. Pericak-Vance and Mayeux). Data generation and harmonization in the Follow-up Phase is supported by U54AG052427 (to Drs. Schellenberg and Wang). The FUS Phase analysis of sequence data is supported through U01AG058589 (to Drs. Destefano, Boerwinkle, De Jager, Fornage, Seshadri, and Wijsman), U01AG058654 (to Drs. Haines, Bush, Farrer, Martin, and Pericak-Vance), U01AG058635 (to Dr. Goate), RF1AG058066 (to Drs. Haines, Pericak-Vance, and Scott), RF1AG057519 (to Drs. Farrer and Jun), R01AG048927 (to Dr. Farrer), and RF1AG054074 (to Drs. Pericak-Vance and Beecham).

The ADGC cohorts include: Adult Changes in Thought (ACT) (UO1 AG006781, UO1 HG004610, UO1 HG006375, U01 HG008657), the Alzheimer’s Disease Centers (ADC) ( P30 AG019610, P30 AG013846, P50 AG008702, P50 AG025688, P50 AG047266, P30 AG010133, P50 AG005146, P50 AG005134, P50 AG016574, P50 AG005138, P30 AG008051, P30 AG013854, P30 AG008017, P30 AG010161, P50 AG047366, P30 AG010129, P50 AG016573, P50 AG016570, P50 AG005131, P50 AG023501, P30 AG035982, P30 AG028383, P30 AG010124, P50 AG005133, P50 AG005142, P30 AG012300, P50 AG005136, P50 AG033514, P50 AG005681, and P50 AG047270), the Chicago Health and Aging Project (CHAP) (R01 AG11101, RC4 AG039085, K23 AG030944), Indianapolis Ibadan (R01 AG009956, P30 AG010133), the Memory and Aging Project (MAP) ( R01 AG17917), Mayo Clinic (MAYO) (R01 AG032990, U01 AG046139, R01 NS080820, RF1 AG051504, P50 AG016574), Mayo Parkinson’s Disease controls (NS039764, NS071674, 5RC2HG005605), University of Miami (R01 AG027944, R01 AG028786, R01 AG019085, IIRG09133827, A2011048), the Multi-Institutional Research in Alzheimer’s Genetic Epidemiology Study (MIRAGE) (R01 AG09029, R01 AG025259), the National Cell Repository for Alzheimer’s Disease (NCRAD) (U24 AG21886), the National Institute on Aging Late Onset Alzheimer's Disease Family Study (NIA- LOAD) (R01 AG041797), the Religious Orders Study (ROS) (P30 AG10161, R01

AG15819), the Texas Alzheimer's Research and Care Consortium (TARCC) (funded by the Darrell K Royal Texas Alzheimer's Initiative), Vanderbilt University/Case Western Reserve University (VAN/CWRU) (R01 AG019757, R01 AG021547, R01 AG027944, R01 AG028786, P01 NS026630, and Alzheimer's Association), the Washington Heights-Inwood Columbia Aging Project (WHICAP) (RF1 AG054023), the University of Washington Families (VA Research Merit Grant, NIA: P50AG005136, R01AG041797, NINDS: R01NS069719), the Columbia University HispanicEstudio Familiar de Influencia Genetica de Alzheimer (EFIGA) (RF1 AG015473), the University of Toronto (UT) (funded by Wellcome Trust, Medical Research Council, Canadian Institutes of Health Research), and Genetic Differences (GD) (R01 AG007584). The CHARGE cohorts are supported in part by National Heart, Lung, and Blood Institute (NHLBI) infrastructure grant HL105756 (Psaty), RC2HL102419 (Boerwinkle) and the neurology working group is supported by the National Institute on Aging (NIA) R01 grant AG033193.

The CHARGE cohorts participating in the ADSP include the following: Austrian Stroke Prevention Study (ASPS), ASPS-Family study, and the Prospective Dementia Registry-Austria (ASPS/PRODEM-Aus), the Atherosclerosis Risk in Communities (ARIC) Study, the Cardiovascular Health Study (CHS), the Erasmus Rucphen Family Study (ERF), the Framingham Heart Study (FHS), and the Rotterdam Study (RS). ASPS is funded by the Austrian Science Fond (FWF) grant number P20545-P05 and P13180 and the Medical University of Graz. The ASPS-Fam is funded by the Austrian Science Fund (FWF) project I904),the EU Joint Programme - Neurodegenerative Disease Research (JPND) in frame of the BRIDGET project (Austria, Ministry of Science) and the Medical University of Graz and the Steiermärkische Krankenanstalten Gesellschaft. PRODEM-Austria is supported by the Austrian Research Promotion agency (FFG) (Project No. 827462) and by the Austrian National Bank (Anniversary Fund, project 15435. ARIC research is carried out as a collaborative study supported by NHLBI contracts (HHSN268201100005C, HHSN268201100006C, HHSN268201100007C, HHSN268201100008C, HHSN268201100009C, HHSN268201100010C, HHSN268201100011C, and HHSN268201100012C). Neurocognitive data in ARIC is collected by U01 2U01HL096812, 2U01HL096814, 2U01HL096899, 2U01HL096902, 2U01HL096917 from the NIH (NHLBI, NINDS, NIA and NIDCD), and with previous brain MRI examinations funded by R01-HL70825 from the NHLBI. CHS research was supported by contracts HHSN268201200036C, HHSN268200800007C, N01HC55222, N01HC85079, N01HC85080, N01HC85081, N01HC85082, N01HC85083, N01HC85086, and grants U01HL080295 and U01HL130114 from the NHLBI with additional contribution from the National Institute of Neurological Disorders and Stroke (NINDS). Additional support was provided by R01AG023629, R01AG15928, and R01AG20098 from the NIA. FHS research is supported by NHLBI contracts N01-HC-25195 and HHSN268201500001I. This study was also supported by additional grants from the NIA (R01s AG054076, AG049607 and AG033040 and NINDS (R01 NS017950). The ERF study as a part of EUROSPAN (European Special Populations Research Network) was supported by European Commission FP6 STRP grant number 018947 (LSHG-CT-2006-01947) and also received funding from the European Community's Seventh Framework Programme (FP7/2007-2013)/grant agreement HEALTH-F4- 2007-201413 by the European Commission under the programme "Quality of Life and Management of the Living Resources" of 5th Framework Programme (no. QLG2-CT-2002- 01254). High-throughput analysis of the ERF data was supported by a joint grant from the Netherlands Organization for Scientific Research and the Russian Foundation for Basic Research (NWO-RFBR 047.017.043). The Rotterdam Study is funded by Erasmus Medical Center and Erasmus University, Rotterdam, the Netherlands Organization for Health Research and Development (ZonMw), the Research Institute for Diseases in the Elderly (RIDE), the Ministry of Education, Culture and Science, the Ministry for Health, Welfare and Sports, the European Commission (DG XII), and the municipality of Rotterdam. Genetic data sets are also supported by the Netherlands

Organization of Scientific Research NWO Investments (175.010.2005.011, 911-03-012), the Genetic Laboratory of the Department of Internal Medicine, Erasmus MC, the Research Institute for Diseases in the Elderly (014-93-015; RIDE2), and the Netherlands Genomics Initiative (NGI)/Netherlands Organization for Scientific Research (NWO) Netherlands Consortium for Healthy Aging (NCHA), project 050-060-810. All studies are grateful to their participants, faculty and staff. The content of these manuscripts is solely the responsibility of the authors and does not necessarily represent the official views of the National Institutes of Health or the U.S. Department of Health and Human Services.

The FUS cohorts include: the Alzheimer's Disease Centers (ADC) ( P30 AG019610, P30 AG013846, P50 AG008702, P50 AG025688, P50 AG047266, P30 AG010133, P50 AG005146, P50 AG005134, P50 AG016574, P50 AG005138, P30 AG008051, P30 AG013854, P30 AG008017, P30 AG010161, P50 AG047366, P30 AG010129, P50 AG016573, P50 AG016570, P50 AG005131, P50 AG023501, P30 AG035982, P30 AG028383, P30 AG010124, P50 AG005133, P50 AG005142, P30 AG012300, P50 AG005136, P50 AG033514, P50 AG005681, and P50 AG047270), Alzheimer's Disease Neuroimaging Initiative (ADNI) (U19AG024904), Amish Protective Variant Study (RF1AG058066), Cache County Study (R01AG11380, R01AG031272, R01AG21136, RF1AG054052), Case Western Reserve University Brain Bank (CWRUBB) (P50AG008012), Case Western Reserve University Rapid Decline (CWRURD) (RF1AG058267, NU38CK000480), CubanAmerican Alzheimer's Disease Initiative (CuAADI) (3U01AG052410), Estudio Familiar de Influencia Genetica en Alzheimer (EFIGA) (5R37AG015473, RF1AG015473, R56AG051876), Genetic and Environmental Risk Factors for Alzheimer Disease Among African Americans Study (GenerAAtions) (2R01AG09029, R01AG025259, 2R01AG048927), Gwangju Alzheimer and Related Dementias Study (GARD) (U01AG062602), Hussman Institute for Human Genomics Brain Bank (HIHGBB) (R01AG027944, Alzheimer's Association "Identification of Rare Variants in Alzheimer Disease"), Ibadan Study of Aging (IBADAN) (5R01AG009956), Mexican Health and Aging Study (MHAS) (R01AG018016), Multi-Institutional Research in Alzheimer's Genetic Epidemiology (MIRAGE) (2R01AG09029, R01AG025259, 2R01AG048927), Northern Manhattan Study (NOMAS) (R01NS29993), Peru Alzheimer's Disease Initiative (PeADI) (RF1AG054074), Puerto Rican 1066 (PR1066) (Wellcome Trust (GR066133/GR080002), European Research Council (340755)), Puerto Rican Alzheimer Disease Initiative (PRADI) (RF1AG054074), Reasons for Geographic and Racial Differences in Stroke (REGARDS) (U01NS041588), Research in African American Alzheimer Disease Initiative (REAAADI) (U01AG052410), Rush Alzheimer's Disease Center (ROSMAP) (P30AG10161, R01AG15819, R01AG17919), University of Miami Brain Endowment Bank (MBB), and University of Miami/Case Western/North Carolina A&T African American (UM/CASE/NCAT) (U01AG052410, R01AG028786).

The four LSACs are: the Human Genome Sequencing Center at the Baylor College of Medicine (U54 HG003273), the Broad Institute Genome Center (U54HG003067), The American Genome Center at the Uniformed Services University of the Health Sciences (U01AG057659), and the Washington University Genome Institute (U54HG003079).

Biological samples and associated phenotypic data used in primary data analyses were stored at Study Investigators institutions, and at the National Cell Repository for Alzheimer's Disease (NCRAD, U24AG021886) at Indiana University funded by NIA. Associated Phenotypic Data used in primary and secondary data analyses were provided by Study Investigators, the NIA funded Alzheimer's Disease Centers (ADCs), and the National Alzheimer's Coordinating Center (NACC, U01AG016976) and the National Institute on Aging Genetics of Alzheimer's Disease Data Storage Site (NIAGADS, U24AG041689) at the University of Pennsylvania, funded by NIA This research was supported in part by the Intramural

Research Program of the National Institutes of health, National Library of Medicine. Contributors to the Genetic Analysis Data included Study Investigators on projects that were individually funded by NIA, and other NIH institutes, and by private U.S. organizations, or foreign governmental or nongovernmental organizations.

### Tables

**Table S1.** Basic demographic information about study participants.

| Dataset | ε2 Analysis | | | | | ε4 Analysis | | | | |
|---|---|---|---|---|---|---|---|---|---|---|
| | N | F% | ε2% | AD% | Age (SD) | N | F% | ε4% | AD% | Age (SD) |
| **ADGC** | 2305 | 58.61 | 14.49 | 53.23 | 74.92 (10.1) | 4460 | 55.25 | 55.81 | 72.31 | 72.43 (9.37) |
| **ADSP-WGS** | 730 | 51.23 | 13.15 | 26.58 | 79.73 (7.90) | 1059 | 50.42 | 40.13 | 39.47 | 78.39 (7.98) |
| **CHS** | 2543 | 60.05 | 17.30 | 4.210 | 83.62 (5.21) | 2701 | 60.16 | 22.14 | 6.550 | 83.41 (5.10) |
| **FHS** | 3159 | 54.67 | 16.43 | 8.040 | 84.04 (8.34) | 3472 | 54.61 | 23.96 | 9.820 | 82.99 (8.58) |
| **LOAD FBS** | 1766 | 62.23 | 13.76 | 31.26 | 77.85 (7.97) | 3367 | 62.55 | 54.77 | 51.71 | 76.49 (7.43) |

Proportions are given in the ε4-negative (ε2 analysis) and ε2-negative (ε4 analysis) samples.

AD = Alzheimer's disease; ADGC = Alzheimer's Disease Centers (ADCs) data from the Alzheimer's Disease Genetics Consortium [1]; ADSP-WGS = whole genome sequencing data from Alzheimer's Disease Sequencing Project [2,3]; CHS = Cardiovascular Health Study [4]; FHS = Framingham Heart Study [5,6]; LOAD FBS = Late Onset Alzheimer's Disease Family Based Study from the National Institute on Aging [7]; F% = percentage of females; AD% = percentage of AD cases; ε2% = percentage of ε2-carriers; ε4% = percentage of ε4-carriers; Age = the average age at onset for AD cases and age at last visit or death for controls; SD = standard deviation.

A smaller percentage of AD cases in FHS and CHS than in the other studies is mainly due to the community-based design of FHS and CHS.

### Figures

**Figure S1.** REACTOME pathways enrichment analysis of genes harboring SNPs in group-specific LD with rs7412.

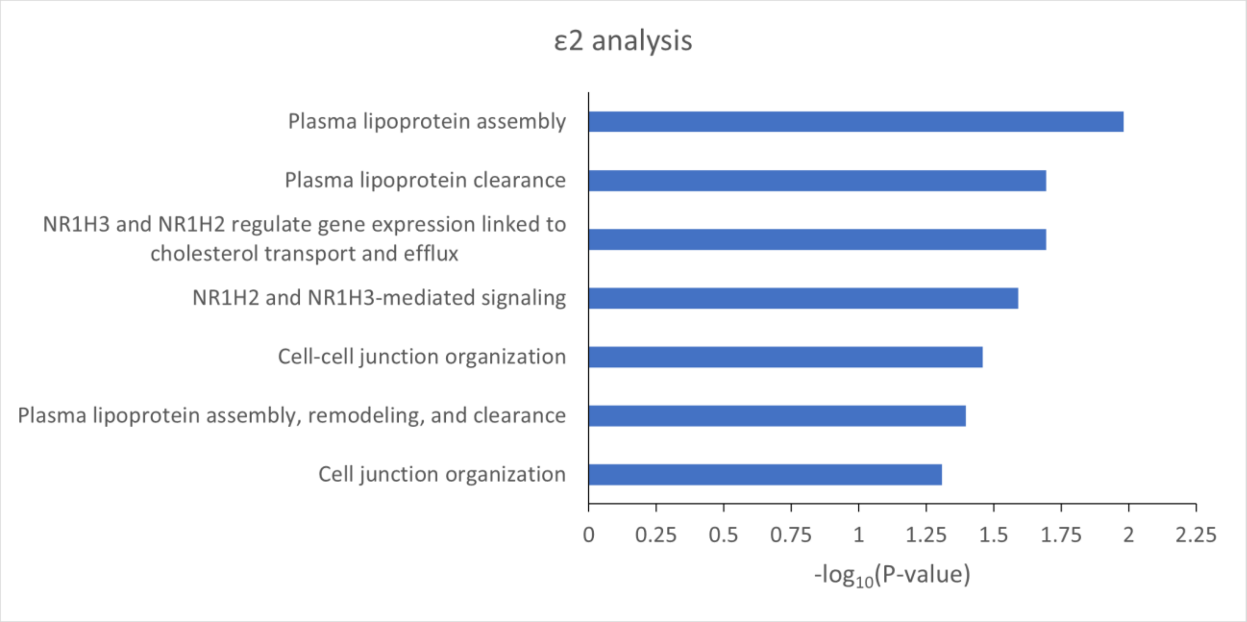

**Figure S2.** REACTOME pathways enrichment analysis of genes harboring SNPs in group-specific LD with rs429358.

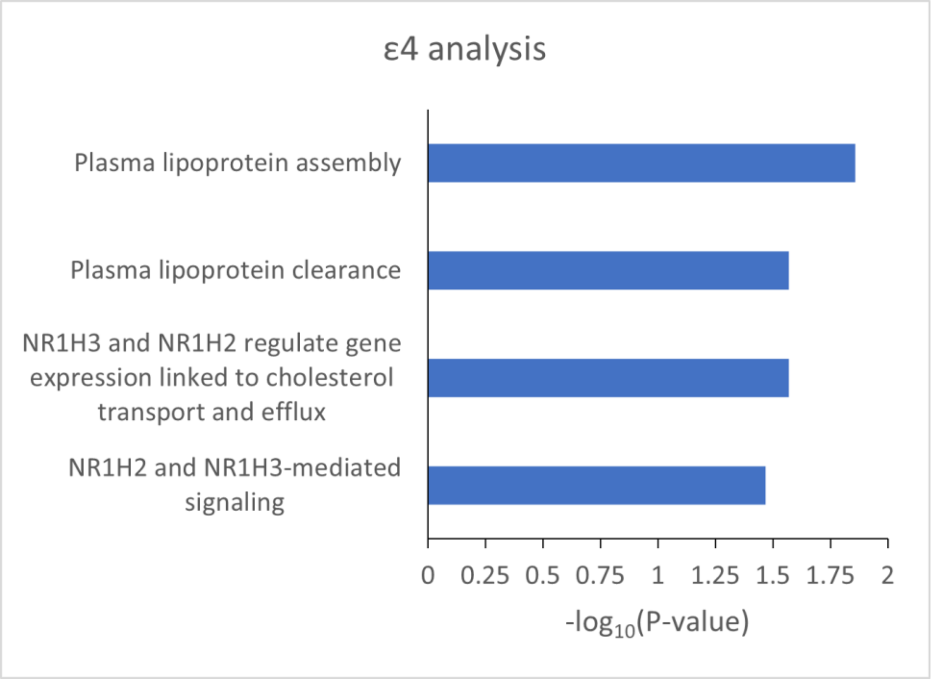

**References**

1. Naj AC, Jun G, Beecham GW, Wang L-S, Vardarajan BN, Buros J, et al. Common variants at MS4A4/MS4A6E, CD2AP, CD33 and EPHA1 are associated with late-onset Alzheimer's disease. Nat Genet. 2011;43:436–41.

2. Beecham GW, Bis JC, Martin ER, Choi S-H, DeStefano AL, van Duijn CM, et al. The Alzheimer's Disease Sequencing Project: study design and sample selection. Neurol Genet. 2017;3:e194.

3. Crane PK, Foroud T, Montine TJ, Larson EB. Alzheimer's Disease Sequencing Project discovery and replication criteria for cases and controls: Data from a community-based prospective cohort study with autopsy follow-up. Alzheimers Dement. 2017;13:1410–3.

4. Fried LP, Borhani NO, Enright P, Furberg CD, Gardin JM, Kronmal RA, et al. The cardiovascular health study: design and rationale. Ann Epidemiol. 1991;1:263–76.

5. Dawber TR, Meadors GF, Moore FE. Epidemiological approaches to heart disease: the Framingham study. Am J Public Health Nations Health. 1951;41:279–86.

6. Feinleib M, Kannel WB, Garrison RJ, McNamara PM, Castelli WP. The Framingham offspring study: design and preliminary data. Prev Med. 1975;4:518–25.

7. Lee JH, Cheng R, Graff-Radford N, Foroud T, Mayeux R. Analyses of the national institute on aging late-onset Alzheimer's disease family study: implication of additional loci. Arch Neurol. 2008;65:1518–26.
